# Acute respiratory distress syndrome and acute kidney injury in critically ill patients: a scoping review protocol about the lung-kidney crosstalk

**DOI:** 10.1101/2024.11.27.24318094

**Authors:** Francisco Z. Mattedi, Heitor S. Ribeiro, Geraldo F. Busatto, Carlos R. R. Carvalho, Emmanuel A. Burdmann, HCFMUSP COVID-19 Study Group

**Affiliations:** LIM 12, Hospital das Clinicas HCFMUSP, Faculdade de Medicina, Universidade de Sao Paulo, São Paulo, BR; Departamento e Instituto de Psiquiatria, Hospital das Clínicas HCFMUSP, Faculdade de Medicina, Universidade de Sao Paulo, São Paulo, BR; Divisão de Pneumologia, Instituto do Coração, Hospital das Clínicas HCFMUSP, Faculdade de Medicina, Universidade de Sao Paulo, São Paulo, BR

**Author notes:** These authors contributed equally and share the first author position. **Corresponding author:** Emmanuel A. Burdmann, MD, PhD. Laboratório de Investigação (LIM) 12, Serviço de Nefrologia, Faculdade de Medicina da Universidade de São Paulo, São Paulo, Brazil, Postal Code: 01246-903.

**Keywords:** acute kidney injury, renal replacement therapy, respiratory distress syndrome, artificial respiration, critically ill

## Abstract

**Introduction:** Acute respiratory distress syndrome (ARDS) is a risk factor for several adverse clinical outcomes, including mortality. Acute kidney injury (AKI) is one of the most common extrapulmonary complications in ARDS. Despite the high incidence of AKI in patients with ARDS, the lung-kidney crosstalk is not yet fully understood.

**Objective:** This scoping review aims to describe the association between ARDS and AKI in critically ill patients.

**Methods:** We will follow the Joanna Briggs Institute methodology and Preferred Reporting Items for Systematic Reviews and Meta-Analyses extension for Scoping Reviews. Based on the **P**articipants, **C**oncept, and **C**ontext framework, we will include studies that investigated critically ill patients with ARDS (**P**articipants), describe AKI-related outcomes (**C**oncept), and were conducted in hospitals (**C**ontext). A comprehensive literature search will be performed using the MEDLINE, Embase, and LILACS databases without date or language restrictions from database inception until January 2024. Observational studies with retrospective, prospective, and case-control designs will be considered. Two independent reviewers will screen titles and abstracts and perform full-text reviews. Data extraction will be performed by the main reviewer and checked by a second one. Data on the diagnosis of ARDS and follow-up time for AKI-related outcomes will be extracted from the selected evidence. The quantitative results will be synthesized and presented in tables and figures along with a narrative summary.

**Ethics and dissemination:** There is no ethical approval requirement for this scoping review. On completion, the review will be published in a peer-reviewed academic journal and presented at a health conference.

## BACKGROUND

Acute respiratory distress syndrome (ARDS) is responsible for 10.4% of admissions to the intensive care unit (ICU) and is a risk factor for several adverse clinical outcomes, including mortality ^1^. Acute kidney injury (AKI) is one of the most common extrapulmonary complications in ARDS ^2,3^. Liu et al. analyzed data from the ARDSNET trial and reported a 24% incidence of AKI in patients with ARDS, which was associated with an increase in mortality (58% vs 28% in patients without AKI) ^4^. McNicholas et al. found an AKI incidence of 39% (15% of mild/moderate cases and 24% of severe cases) in patients with ARDS in the LUNG SAFE study cohort. AKI was associated with high mortality in both the ICU and hospital settings, and the mortality rates varied according to the AKI severity (49.9% in the mild/moderate AKI group and 57.9% in the severe group, compared with 30.8% in the group without AKI). Patients with ARDS-associated AKI had a longer hospital stay and fewer days free from mechanical ventilation (MV) compared with patients without AKI ^2^.

Despite the high incidence of AKI in patients with ARDS, the lung-kidney crosstalk is not yet fully understood. ARDS-associated AKI may be a consequence of the critical illness context and develop independently, contributing to a higher mortality risk. On the other hand, ARDS may act as an independent risk factor for the development of AKI, participating in its pathophysiology, as suggested by previous studies ^3,4^. The lung-kidney crosstalk might occur in both directions, with acute lung injury (ALI) leading to AKI and vice versa. Even so, this crosstalk remains to be explored, and investigations of temporal association, the impact on kidney function recovery, persistence of kidney replacement therapy (KRT), and the progression to chronic kidney disease are lacking.

The possible mechanisms of the lung-kidney crosstalk are related to hemodynamic, neurohormonal, and inflammatory changes caused by a primary lung injury. In a cohort of patients with ARDS, increased levels of interleukin (IL)-6, soluble tumor necrosis factor receptors (sTNFR-I and -II), and plasminogen activator inhibitor-1 (PAI-1) were markedly associated with AKI development ^4^. Hypoxemia and hypercapnia are associated with increased renal vascular resistance ^5,6^. Positive end-expiratory pressure (PEEP) during MV was related to a reduction in cardiac output, glomerular filtration rate (GFR), and renal blood flow, in addition to the activation of antidiuretic hormone and the renin-angiotensin-aldosterone system, promoting water retention ^7^.

Biotrauma induced by MV is associated with an increase in the levels of inflammatory markers, including IL-1-beta, IL-6, IL-8, and TNF-alpha, mainly in patients undergoing non-protective ventilation. There are some data showing an increased risk of AKI development after ARDS, regardless of the ventilation parameters used ^8,9^. In a rabbit model of ALI, the animals were randomized to receive protective or conventional (non-protective) ventilation ^10^. The conventional ventilation group showed an increase in the levels of inflammatory markers and serum creatinine (sCr) and apoptosis compared with the protective ventilation group. In a similar model in pigs, infiltration of inflammatory cells in the kidney, heart, and liver was noted among animals undergoing conventional ventilation compared with the group receiving protective ventilation ^11^.

Data on AKI after ARDS in the clinical setting are not consistent, and there is high heterogeneity in the definitions of ARDS and AKI. ARDS is mainly defined by the Berlin criteria ^12^; however, the American-European Consensus Conference (AECC)^13^ criteria is also used. For AKI, definitions based on the Kidney Disease: Improving Global Outcomes (KDIGO) 2012; Acute Kidney Injury Network; and Risk, Injury, Failure, Loss of Kidney Function, and End-stage Kidney Disease criteria and other adapted definitions have been used. Outcomes are also heterogeneous. The most frequently evaluated outcomes in patients with ARDS are the incidence of AKI and mortality. However, the time to develop AKI, incidence of KRT, recovery of renal function, and progression to CKD are not reported in most articles. Hence, the impact of ARDS as a risk factor for AKI remains poorly evaluated.

Darmon et al. demonstrated that 44.3% of patients with ARDS admitted to the ICU developed AKI compared with 27.4% of patients without ARDS. Mortality was also higher in the ARDS-AKI group than in the ARDS group without AKI (27.6% vs 8.1%). In this study, ARDS was identified as an independent risk factor for AKI (OR, 11.01; 95% CI, 6.83 to 17.73) ^3^. A Brazilian study that prospectively evaluated 100 patients admitted to the ICU with pulmonary disease who subsequently developed ARDS showed that a partial pressure of oxygen in arterial blood/fraction of inspiratory oxygen concentration (PaO_2_/FIO_2_, P/F ratio) lower than 200 mmHg on admission was independently associated with subsequent AKI development ^14^. Clemens et al. evaluated 830 patients admitted to the USAISR Burn Center and showed an AKI incidence of 27% in patients who developed ARDS. In univariate analysis, ARDS was identified as an independent risk factor for AKI; however, this result was not confirmed in the multivariate analysis ^15^. In a retrospective cohort of patients hospitalized with coronavirus disease 2019 (COVID-19), the incidence of AKI was higher in the group with ARDS/MV (89.7% vs. 21.7% in non-ventilated patients)^16^. In a recent meta-analysis, a higher incidence of AKI and the need for KRT was found in COVID-19 cohorts with ARDS compared with those without it ^17^. ARDS was not described as an independent risk factor for AKI in the KDIGO 2012 guidelines and the KDIGO Controversies Conference on Acute Kidney Injury in 2019 ^18,19^.

A systematic review and meta-analysis of 17 studies evaluating the incidence and clinical outcomes in patients who developed AKI after ARDS was recently published ^20^. Therein, the pooled incidence rate of AKI after ARDS was 39% (95% CI, 32 to 46%). Furthermore, the mortality rate was higher in the patients who developed AKI after ARDS. ARDS was an independent risk factor for the development of AKI in only two studies. Although these results indicate a possible association between ARDS and AKI, prospective cohort studies specifically designed to answer this question are needed to improve the understanding of the ARDS-AKI association.

Considering the heterogeneity of clinical data regarding lung-kidney crosstalk, we decided to conduct this scoping review to evaluate the evidence on the relationship between these two clinical entities and the gaps present in the literature.

### Review question

How is the association between ARDS and AKI described in the literature?

## ELIGIBILITY CRITERIA

Eligibility criteria based on the PCC framework are shown in **Table 1** and described below in detail.

**Table 1.** Participants, Concept, Context (PCC) framework.

|  | Definition |
| --- | --- |
| **Participants** | Adults with acute lung injury/ARDS |
| **Concept** | AKI incidence, AKI stages, need/use for/of KRT, and kidney function recovery |
| **Context** | ICU, ward, emergency room, and community settings |

AKI, acute kidney injury; ICU, intensive care unit; KRT, kidney replacement therapy.

### Participants

In this scoping review, adults (aged ≥18 years) who were in the ICU, ward, or emergency room with ALI/ARDS will be considered. The last group will be diagnosed based on the most prominent guidelines, such as the Berlin criteria^12^ and the AECC criteria^13^. However, studies that applied other definitions will also be considered.

### Concept

We will use sources pertaining to the assessment of patients with ALI/ARDS, which also included the assessment of kidney function (sCr and/or urinary output), diagnosis of AKI (stages of AKI), time to the diagnosis, description of the need for or use of KRT, and kidney function recovery. AKI will be defined based on the KDIGO^18^, AKIN^21^, and RIFLE^22^ criteria or the use of KRT. However, studies that adopted other AKI definitions will also be considered.

### Context

Evidence from investigations of ARDS and kidney function conducted in the ICU, ward, emergency room, and community settings will be considered.

### Types of Sources

All types of observational study designs (e.g., retrospective, prospective, and case-control studies) will be considered. Reviews, commentaries, and letters to the editor will not be included.

## METHODS

The proposed scoping review will be conducted following the Joanna Briggs Institute (JBI) methodology for scoping reviews^23^ and the Preferred Reporting Items for Systematic Reviews and Meta-Analyses extension for Scoping Reviews (PRISMA-ScR)^24^.

### Search Strategy

We designed a search strategy to mainly locate published evidence by a peer-review process. A preliminary search on MEDLINE was undertaken to identify relevant articles on the topic. The text words contained in the titles and abstracts of the relevant articles and the index terms used to describe the articles were used to develop a full search strategy for the MEDLINE, EMBASE, and LILACS databases. The search strategy, including all identified keywords and index terms, has been adapted for each database and/or information source (see **Appendix 1**). The databases were searched from inception to January 2024. The reference lists of all the included sources of evidence will be screened for additional studies; no language restrictions will be applied. Sources of unpublished studies and gray literature to be searched include Google Scholar and Google up to the fifth search page.

### Source of evidence selection

All identified citations were collated and uploaded into the Covidence systematic review software (Veritas Health Innovation, Melbourne, AU), where duplicates were removed for the screening process. Following a pilot test, the titles and abstracts will be screened by two independent reviewers (FZM and HSR) for assessment against the inclusion criteria. The full text of the selected evidence will be assessed in detail by the same independent reviewers. Reasons for the exclusion of sources of evidence in full texts that do not meet the inclusion criteria will be recorded. Any disagreements between the reviewers at each stage of the selection process will be resolved through discussion or by consulting an additional reviewer (EAB). The results of the search and the study inclusion process will be reported in full in the final scoping review and presented in the PRISMA-ScR flow diagram.

### Data extraction

Data will be extracted from the studies included in the scoping review by the main reviewer (FZM) and checked by a second reviewer (HSR) by using a data extraction spreadsheet adapted from the JBI Manual for Evidence Synthesis ^25^. The extracted data will include specific details about MV, diagnosis of ARDS, kidney function, development of AKI, and other items. Additional general information will be extracted, including the author(s), year of publication, and country. A draft extraction form is provided (see **Appendix 2**). The draft data extraction tool will be modified and revised as necessary during the process of data extraction. Any disagreements between the reviewers will be resolved through discussion or by consulting an additional reviewer (EAB). If necessary, the authors of the studies will be contacted to request missing or additional data.

### Quality assessment

Methodological quality assessment of the included studies will be conducted independently by two reviewers (FZM and HSR) by using the JBI Critical Appraisal Checklist^26^. The JBI tool consists of nine items with four possible responses (*Yes, No, Unclear*, or *Not applicable*). The greater the number of *Yes* answers, the better the quality of the study. We will dichotomize the studies high quality (below the median) and low quality (above the median) for further analysis. Disagreements will be resolved through consensus or by consulting a third reviewer (EAB).

### Data analysis and presentation

The findings from the scoping review will be reported per the PRISMA-ScR guideline^24^. Data will be synthesized and presented in tables and figures along with a narrative summary. Statistical analysis will be performed using the Statistical Package for the Social Sciences (version 29.0, IBM Corp., Armonk, NY, USA).

### Patient and public involvement

There will be no patient and public involvement during the review process; however, results will be shared and disseminated throughout an extended network within the Hospital das Clínicas da Universidade de São Paulo (HCFMUSP), which includes patients, their relatives, stakeholders, and others.

### Ethics and dissemination

There is no requirement for ethical approval for this scoping review. On completion, it will be published in a peer-reviewed academic journal and presented at a conference.

## Data Availability

The datasets to be analyzed during the current study will be available from the corresponding author upon reasonable request.

## Acknowledgments

We would like to thank Editage (www.editage.com) for English language editing.

## Authors’ contributions

FZM, HSR, and EAB conceived and designed the study. FZM wrote the first draft. HSR, GFB, CRRC, and EAB critically reviewed the manuscript. All authors read and approved the final version.

## Appendix I Search strategy

### MEDLINE – 352 references (18th January 2024)

#1 “Respiratory Distress Syndrome”[MeSH Terms] OR “Respiratory Insufficiency”[MeSH Terms] OR “respiration, artificial”[MeSH Terms] OR “Acute Lung Injury”[MeSH Terms] OR “Acute Chest Syndrome”[MeSH Terms]

#2 “Acute Kidney Injury”[MeSH Terms] OR “AKI”[All Fields] OR “AKF”[All Fields]

#3 “Intensive Care Units”[MeSH Terms] OR “Emergency Medical Services”[MeSH Terms] OR “Community Health Services”[MeSH Terms] OR “emergency service, hospital”[MeSH Terms] OR “Critical Care”[MeSH Terms]

#4 #1 AND #2 AND #3

#5 NOT animal*

#6 #4 AND #5

#7 NOT “child*”[All Fields] OR “infant*”[All Fields] OR “pediatric*”[All Fields]

#8 #6 AND #7

#### Embase – 2548 references (18th January 2024)

#1 ‘adult respiratory distress syndrome’ OR ‘acute lung injury’ OR ‘invasive ventilation’ OR ‘noninvasive ventilation’

#2 acute AND kidney AND injury

#3 ‘acute kidney failure’ OR AKI OR AKF

#4 #2 OR #3

#5 ‘intensive care unit’ OR ‘emergency ward’ OR ‘community care’

#6 emergency AND care AND unit

#7 critical AND care AND unit

#8 #5 OR #6 OR #7

#9 #1 AND #4 AND #8

#10 NOT animal*

#11 #9 AND #10

#12 NOT child* NOT infant* NOT pediatric*

#13 #11 AND #12

#### LILACS – 43 references (18th January 2024)

Síndrome do Desconforto Respiratório OR Síndrome de Dificultad Respiratoria OR acute respiratory distress syndrome AND Injúria Renal Aguda OR Lesión Renal Aguda OR acute kidney injury

## Appendix II Draft extraction

|  | Characteristics |  |  |  |  |  |  |  |  |  |  |
| --- | --- | --- | --- | --- | --- | --- | --- | --- | --- | --- | --- |
| Author | Year | Country | Region | Study design | Sample characteristic | Sample | Men | Woman | Age (years) | AKI incidence in ARDS patients | Severity tool |

| No ARDS | ARDS | Diagnostic criteria for ARDS | ARDS definition | Mild - P/F 200-300 | Moderate - P/F 100-200 | Severe - P/F < 100 |
| --- | --- | --- | --- | --- | --- | --- |

| AKI Criteria | AKI biomarker | AKI definition | AKI diagnosis (time) | AKI with mild ARDS | AKI with moderate ARDS | AKI with severe ARDS | AKI 1 (risk) | AKI 2 (injury) | AKI 3 (failure) | Risk factors |
| --- | --- | --- | --- | --- | --- | --- | --- | --- | --- | --- |

| Outcomes |  |  |  |  |  |
| --- | --- | --- | --- | --- | --- |
| ICU lenght of stay | Hospital lenght of stay | ICU mortality | Hospital Mortality | AKI recovery | Dialysis |

